# Susceptibility Signatures of the Basal Ganglia and Hippocampal Formation in Acute Mild Traumatic Brain Injury: A Quantitative Susceptibility Mapping Study

**DOI:** 10.1101/2025.01.09.25320291

**Authors:** Christi A. Essex, Mayan J. Bedggood, Jenna L. Merenstein, Helen C. Murray, Catherine Morgan, Samantha J. Holdsworth, Richard L. M. Faull, Patria Hume, Alice Theadom, Mangor Pedersen

**Affiliations:** Department of Psychology and Neuroscience, Auckland University of Technology, Auckland 0627, New Zealand; Department of Anatomy and Medical Imaging, Faculty of Medical and Health Sciences, The University of Auckland, Auckland 1023, New Zealand; Center for Brain Research, The University of Auckland, Auckland 1023, New Zealand; Brain Imaging and Analysis Center, Duke University Medical Center, Durham, NC 27710, United States; Center for Advanced MRI, The University of Auckland, Auckland 1023, New Zealand; School of Psychology, The University of Auckland, Auckland 1023, New Zealand; Matai Medical Research Institute, Gisborne 4010, New Zealand; Sports Performance Research Institute New Zealand, Auckland University of Technology, Auckland 0627, New Zealand

**Keywords:** basal ganglia, brain iron, concussion, hippocampal formation, mild traumatic brain injury, quantitative susceptibility mapping

## Abstract

Despite vulnerability to microstructural tissue damage following mild traumatic brain injury (mTBI), key subcortical brain regions have been overlooked in quantitative susceptibility mapping (QSM) studies. Alterations to tissue composition in the functionally and structurally distinct hippocampal subregions, basal ganglia, and associated nuclei may reflect distinct symptomatology, and better characterisation of these regions is needed to increase our understanding of mTBI pathophysiology. We analysed differences in net positive and negative QSM values between 25 males with acute (*<* 14 days) sports-related mTBI (sr-mTBI) and 25 age-matched male controls across 10 hippocampal subregions and 16 basal ganglia and associated nuclei. Additional variables of interest including age, injury severity, and days since injury at the time of magnetic resonance imaging (MRI) were also correlated with both positive and negative susceptibility values. Primary analyses indicated no significant difference in positive susceptibility values between sr-mTBI participants and controls for hippocampal and basal ganglia ROIs. For negative sign values, susceptibility was significantly less negative for sr-mTBI participants in the cornu ammonis 4 (CA4) subfield only (*q* = 0.04). In line with the known linear relationship between iron deposition and age in deep grey matter, particularly within the first three decades of life, significant positive relationships were observed between net positive susceptibility and age in the putamen, caudate, red nucleus, parabrachial pigmented nucleus, and ventral pallidum (*q <* 0.05). Positive relationships were also observed between absolute negative susceptibility values and age in the hippocampal fimbria, caudate, and extended amygdala (*q <* 0.05), suggesting age-related calcifications in these regions. A negative relationship was observed between absolute negative values and age in the ventral pallidum (*q* = 0.04), indicating potential changes to myelin content in this region. No significant associations were observed between any other variable and signed susceptibility values. The results of this study contribute to, and extend, prior literature regarding the temporal kinetics of biomagnetic substrates as a function of ageing. Decreased negative susceptibility after mTBI in the CA4 subfield also suggests potential injury-related effects on myelin content or neuron loss; a particularly interesting finding in light of the well-established vulnerability of cell populations in this region and susceptibility to pathology in chronic traumatic encephalopathy (CTE). The lack of other significant between-group differences suggest that alterations to tissue content may not be quantifiable at the acute stage of injury in subcortical ROIs or may be masked by age-related tissue susceptibility changes as a common feature across all participants in this young cohort. Future research should consider the use of longitudinal study designs to mitigate the influence of these factors.

## Introduction

Whilst participation in sports can have many physical and psychological benefits, there is a non-negligible risk of sustaining a mild traumatic brain injury (mTBI). This is the most common form of traumatic brain injury (TBI),^1^ and reports suggest that up to 30% of all TBI cases are mild head injuries resulting from sports participation.^2^ The related changes to cognition, mental health, and physiology can be far from what categorisation as a “mild” injury would suggest, significantly degrading quality of life and participation in activities of daily living.^3^ These symptoms are generally associated with the acute and subacute stages, but may persist for decades after injury.^4–6^ In addition, repeated instances of mild head trauma expose individuals the risk of developing a neurodegenerative condition, chronic traumatic encephalopathy (CTE),^3,7^ and increase the risk of premature mortality.^8^ Given the popularity of sports the world over, the issue of mTBI warrants careful consideration.

A mTBI is induced by either impact or inertial forces^9^ which transmit mechanical energy to the brain^10^ causing primary and secondary injury.^11^ Primary injury occurs at the time of the incident when the brain is displaced within the skull, damaging the neurons, glia, and blood vessels of the cerebral tissues.^12^ mTBI is most often associated with a diffuse primary injury, specifically diffuse axonal injury, whereby the cellular nerve fibers are stretched, causing cytoskeletal damage.^11,13,14^ Secondary injury refers to the resultant dynamic physiological processes catalysed by the primary injury; this complex and self-perpetuating cascade of biomechanical and metabolic disturbance within the affected tissues can propagate for months after the initial impact.^15^ This includes, but is by no means limited to, altered neurotransmitter signalling, changes to cerebral blood flow, mitochondrial dysfunction, blood-brain barrier (BBB) disruption, oxidative stress, demyelination, and neuroinflammatory responses.^9,11,16^ However, the underlying pathophysiology of mTBI remains poorly understood.^14^

Iron is an essential co-factor for many neuronal processes including mitochondrial respiration, oxygen transport, cellular metabolism, and synthesis of neurotransmitters, myelin, and DNA.^17,18^ However, iron overload, particularly in non-haem form (i.e., iron bound to proteins like ferritin and transferrin rather than haem groups), is increasingly recognised as a driver of secondary neurotoxic events^11,19^ including oxidative stress, cellular degeneration, and inflammation.^20–22^ Non-haem iron is also the primary source of paramagnetic signals assessed by quantitative susceptibility mapping (QSM), an analytical technique of magnetic resonance imaging (MRI) data that enables quantification of biomagnetic substances in neural tissue.^23–26^ As such, mapping distributions of positive susceptibility across the brain as a proxy for iron load represents a promising avenue for investigating potential biomarkers of neuropathology following mTBI. Myelin constitutes the primary source of diamagnetic contrast on QSM,^27^ but calcium^28–30^ and certain proteins such as beta-amyloid (Aβ) and tau also contribute^31–33^ and concentrations of all aforementioned biomagnetic substrates may change as a function of injury-related processes in mTBI.^11,16,22,34^ Isolation of these susceptibility sources may therefore provide crucial insight into the subtle biodynamics and injury cascades occurring shortly after injury.

Research using QSM to examine alterations to grey matter after mTBI have focused on markers of iron accumulation in the basal ganglia as a proxy for secondary injury processes. Iron, as a fundamental component of neurotransmitter synthesis, is more abundant in these deep regions compared to cortical grey matter^35^ supporting the high metabolic demand related to signal processing.^11^ Dense concentrations of iron are particularly noteworthy in the globus pallidus, red nucleus, substantia nigra, putamen, caudate, and thalamus^35^ and may predispose these regions to iron-mediated disorders.^36^ Additional vulnerability to shear and strain damage in mTBI^37^ due to axons originating from, terminating in, or intersecting with these nuclei,^11,38,39^ for example thalamo-cortical projections,^40^ may further increase the risk of iron-driven secondary injury.

Within this existing body of research, a focus on either total deep grey matter,^41,42^ basal ganglia segmentations,^43–45^ or a combination of the two,^46,47^ have predominated. Where segmentations have been employed, the analyses are typically confined to major subcortical structures such as the caudate, putamen, thalamus, and globus pallidus,^43,44,46^ potentially overlooking the susceptibility of many other deep grey matter regions to cytotrauma. To date, only two studies^45,47^ have extended their investigations to include additional subcortical structures such as the nucleus accumbens and amygdala, and the allocortical hippocampus, which exhibit similar iron-related vulnerability as the basal ganglia. However, even these more comprehensive examinations have lacked the anatomical specificity needed to identify distinct structural and functional subregions that may be differentially affected by mTBI, representing a crucial oversight in the QSM-mTBI literature.^11^ These include distinct pallidal substructures, limbic structures, and other associated nuclei. Examining these additional subregions may be critical for identifying potentially more subtle mTBI-related changes to biomagnetic substrates that could otherwise be washed out when examining entire regionsof-interest (ROI), as we have previously demonstrated in the cortical grey matter.^48^

Pathological changes in these deep brain structures responsible for motor control,^49,50^ emotional regulation,^51^ learning,^52^ and memory,^53^ may also drive the heterogeneity of mTBI symptomatology. Iron overload in the deep grey matter in particular may significantly alter cognitive function after mTBI.^54^ The convergent evidence indicates that more comprehensive assessments of the subcortical nuclei are warranted. In addition, examining the hippocampus using a whole-ROI approach neglects the structural and functional distinctions between its subregions. The hippocampus is particularly vulnerable to injury,^55^ and as a central hub for memory^56^ damage to this area may be related to the memory impairments characteristic of mTBI.^57^ Macroscopic methods may thus risk masking specific hippocampal pathologies that are likely related to mTBI symptomatology. In addition, temporal regions, including the hippocampus, also exhibit region-specific atrophy and tau hyperphosphorylation (p-tau) in CTE,^58–60^ which may be related to iron dyshomeostasis.^22,61,62^

Taken together, these studies suggest that investigations differentiating between distinct brain regions and their substructures are necessary to better understand the pathology of mTBI, and may assist in elucidating the genesis of specific injury-related deficits. To contribute to the sparse extant literature, we conducted the first QSM analysis of mTBI effects in the hippocampal subregions alongside the most detailed segmentation of the subcortical nuclei to date. This study aimed to: 1) assess the effects of mTBI on positive (iron-related) and negative (myelin-, protein-, calcium-related) net magnetic susceptibility to better understand acute pathology in, a) 16 segmentations of the basal ganglia and extended nuclei, and b) 10 segmentations of the hippocampus, and; 2) elucidate the relationship between magnetic susceptibility in these regions and potentially moderating variables including age, and injury latency and severity. Although many of these regions are both high in iron and vulnerable to mTBI-related pathology, the limited prior literature has produced mixed results regarding the direction of iron-related effects, if any. Furthermore, the absence of QSM thresholded for inter-voxel sign in previous studies precludes the development of robust hypotheses regarding the directionality of negative susceptibility effects in grey matter. As such, there were no specific *a priori* hypotheses related to either susceptibility sign, nor secondary correlational analyses.

## Materials and Methods

Ethical approval for this research was obtained from the Health and Disabilities Ethics Committee (HDEC) (Date: 18/02/2022, Reference: 2022 EXP 11078) and institutional approval was also obtained from the Auckland University of Technology Ethics Committee (AUTEC) (Date: 18/02/2022, Reference: 22/12). In accordance with the Declaration of Helsinki, all participants provided written informed consent prior to data collection.

### Participants

Data from 25 male contact sports players (*M* = 21.10 years old [16-32], *SD* = 4.59) with acute sports-related mTBI (sr-mTBI) (< 14 days; *M* = 10.40 days, *SD* = 3.03) and 25 age-matched male controls (*M* = 21.10 years old [1632], *SD* = 4.35) were used for this observational study (see Table 1). To mitigate any potential confounding effects due to the known linear relationship between brain iron content and age,^17,35,63–67^ particularly in this young cohort, we ensured that ages were not significantly different between groups (*t*(48) = 0.00, *p* = 1.00). Clinical sr-mTBI participants were recruited through three Axis Sports Medicine clinics (Auckland, New Zealand), via print and social media advertisements, word-of-mouth, and through community-based pathways including referrals from healthcare professionals and sports team management. Each clinical participant was required to have a confirmed sr-mTBI diagnosis by a licensed physician as a prerequisite for study inclusion, and symptom severity was assessed using the Brain Injury Screening Tool (BIST)^68^ either upon presentation to Axis clinics or electronically following recruitment. Healthy controls (HC) were recruited through print and social media advertisements, and word-of-mouth. Exclusion criteria for all participants included a history of significant medical or neurological conditions unrelated to the study’s objectives and contraindications for MRI. Additionally, controls were excluded if they had any recent history of mTBI events (< 12 months) or were living with any long-term effects of previous mTBI. All participants completed a brief demographic questionnaire and attended a 1-hour MRI scan at The Centre for Advanced MRI (CAMRI), Auckland, New Zealand. All scans were reviewed by a certified neuroradiologist consultant for clinically significant findings. While some incidental findings were identified, none were considered to be clinically significant and required no follow-up. MRI findings for this cohort are reported in extensive detail in previous publications.^48^

**Table 1:** Summary of sr-mTBI participant clinical characteristics.

| ID | Age | DSI | BISTScore | MOI |
| --- | --- | --- | --- | --- |
| mTBI-01 | < 20 | 5 days | 140 | Rugby |
| mTBI-02 | < 20 | 5 days | 12 | Rugby |
| mTBI-03 | 20s | 6 days | 78 | Rugby |
| mTBI-04 | < 20 | 13 days | 18 | Rugby |
| mTBI-05 | < 20 | 12 days | 61 | Rugby |
| mTBI-06 | 20s | 13 days | 42 | Rugby |
| mTBI-07 | 20s | 13 days | 13 | Football |
| mTBI-08 | 20s | 12 days | 6 | Hockey |
| mTBI-09 | 20s | 6 days | 56 | Rugby |
| mTBI-10 | < 20 | 12 days | 54 | Rugby |
| mTBI-11 | 20s | 10 days | 52 | Rugby |
| mTBI-12 | 30s | 13 days | 13 | Football |
| mTBI-13 | < 20 | 5 days | 79 | Rugby |
| mTBI-14 | 20s | 13 days | 2 | Rugby |
| mTBI-15 | < 20 | 13 days | 22 | Rugby |
| mTBI-16 | < 20 | 8 days | 117 | Futsal |
| mTBI-17 | 20s | 13 days | * | Rugby |
| mTBI-18 | 20s | 10 days | 34 | Gymnastics |
| mTBI-19 | 20s | 13 days | 28 | Jiu-jitsu |
| mTBI-20 | 20s | 11 days | 69 | Surfing |
| mTBI-21 | < 20 | 7 days | 14 | Rugby |
| mTBI-22 | < 20 | 13 days | 39 | Judo |
| mTBI-23 | < 20 | 9 days | 34 | Rugby |
| mTBI-24 | < 20 | 12 days | 68 | Rugby |
| mTBI-25 | 20s | 12 days | 17 | Rugby |
| Mean | 21.10 (4.59) years | 10.4 (3.03) days | 44.5 (35.0) /160 |  |

*Note.* Diagnostic assessment was limited to the volume T1, SWI and DWI sequences with only limited interpretation of the multi-echo T2 stack. Clinical assessments were relevant to the identification of micro-haemorrhages, areas of siderosis, T1 appearance, gliosis, volume, ventricular volumes and non-neurological findings. Age is reported in a range to prevent re-identification of participants. The possible range of BIST scores is 0 (min) to 160 (max). Clinical group data corresponded to date at MRI only with the exception of the BIST<sup>68</sup> acquired >24 hours post-injury prior to MRI scanning (<14 days post). Abbreviations are: mTBI = mild traumatic brain injury; HC = healthy control; ID = unique identifier; DSI = days since injury; BIST = Brain Injury Screening Tool<sup>68</sup>; MOI = mechanism of injury; MRI = magnetic resonance imaging; L = Left; R = right; \* = missing data (BIST incomplete on the Axis Sport Medicine Clinic patient portal, reason unknown).

### Neuroimaging

Details on image acquisition and processing have been previously reported,^48^ and are summarised here for brevity.

#### Acquisition

MRI data were acquired on a 3T Siemens MAGNETOM Vida Fit scanner (Siemens Healthcare, Erlangen, Germany) equipped with a 20-channel head coil. A 3D flow-compensated single-echo gradient-recalled echo (GRE) sequence was used to obtain magnitude and unfiltered phase images suitable for QSM reconstruction. Data were collected at 1 mm isotropic voxel size with matrix size = 180 x 224 x 160 mm, TR = 30 ms; TE = 20 ms; FA = 15^◦^; FoV = 180 mm (LR) × 224 mm (AP) in a total acquisition time of ∼3.43 minutes. For each participant, a high-resolution 3D T_1_weighted (T_1_w) anatomical image volume was acquired for coregistration and segmentation using a MagnetisationPrepared Rapid Acquisition Gradient Echo (MPRAGE) sequence (TR = 1940.0 ms; TE = 2.49 ms, FA = 9^◦^; slice thickness = 0.9 mm; FoV = 230 mm; matrix size = 192 x 512 x 512 mm; GRAPPA = 2; voxel size 0.45 x 0.45 x 0.90 mm) for a total acquisition time of ∼4.31 minutes. DICOMs were converted to NIfTI files and transformed to brain imaging data structure (BIDS)^69^ for further processing using Dcm2Bids^70^ version 3.1.1, which is a wrapper for dcm2niix^71^ (v1.0.20230411).

#### QSM processing

QSM images were reconstructed using QSMxT^72^ v6.4.2 and used a robust two-pass combination method for artefact reduction,^73^ rapid open-source minimum spanning tree algorithm (ROMEO) for phase unwrapping,^74^ background field removal with projection onto dipole fields (PDF),^75^ sparsity-based rapid two-step dipole inversion (RTS),^76^ and whole-brain susceptibility referencing; a pipeline congruent with recent consensus statement recommendations for best-practice QSM reconstruction.^77^ For each participant, the raw magnitude image was skull-stripped using FSL’s *BET*^78^ with robust brain centre estimation and a fractional intensity threshold of between 0.3 and 0.6. Binary masks were derived from the skullstripped magnitude image and applied to the susceptibility maps to erode non-brain signal around the brain perimeter using *fslmaths*. Skull-stripped magnitude images were linearly registered to the California Institute of Technology’s 168 (CIT168) T_1_w template^79^ in Montreal Neurological Institute 152 (MNI152) space using FMRIB’s Linear Transformation Tool (FLIRT)^80–82^ with 12 degrees of freedom (DoF) suitable for atlas-based registration. The resulting transformation matrix was then used for spatial normalisation of the QSM images using *FLIRT*.^80–82^ Using inter-voxel thresholding methods,^83,84^ QSM maps were then thresholded into separate maps of net positive voxels (*QSM^+^*) and net negative (*QSM^−^*) voxels with *fslmaths*, by separating values across voxel boundaries above (*QSM^+^*) and below (*QSM*^−^) zero. This approach may enable more targeted analyses of specific susceptibility sources without the confounding effects of inter-voxel averaging. Both *QSM^+^* and *QSM^−^* susceptibility maps were used in subsequent analyses.

#### Basal ganglia segmentation

The CIT168 basal ganglia mask, available in MNI152 space, provides a detailed 16-part probabilistic atlas of the basal ganglia and associated structures.^79^ Derived from T_1_w and T_2_-weighted in vivo structural images of 168 participants from the Human Connectome Project,^85^ this atlas is openly accessible via the NeuroVault Collection (No. 3145). The segmentations included: the striatum, comprised of the putamen (Pu), caudate (Ca), and nucleus accumbens (NAC); the pallidum, which includes the globus pallidus externus (GPe), globus pallidus internus (GPi), and ventral pallidum (VeP); and the substantia nigra, including the substantia nigra pars compacta (SNc) and substantia nigra pars reticulata (SNr). Additional limbic and hypothalamic structures included the extended amygdala (EXA), hypothalamus (HTH) and mammillary nucleus (MN), the mesolimbic ventral tegmental area (VTA) and the associated parabrachial pigmented nucleus (PBP), the epithalamic habenular nuclei (HN), the subthalamic nucleus (STH), and the red nucleus (RN) (see Fig. 1 for reference).

**Fig 1:**
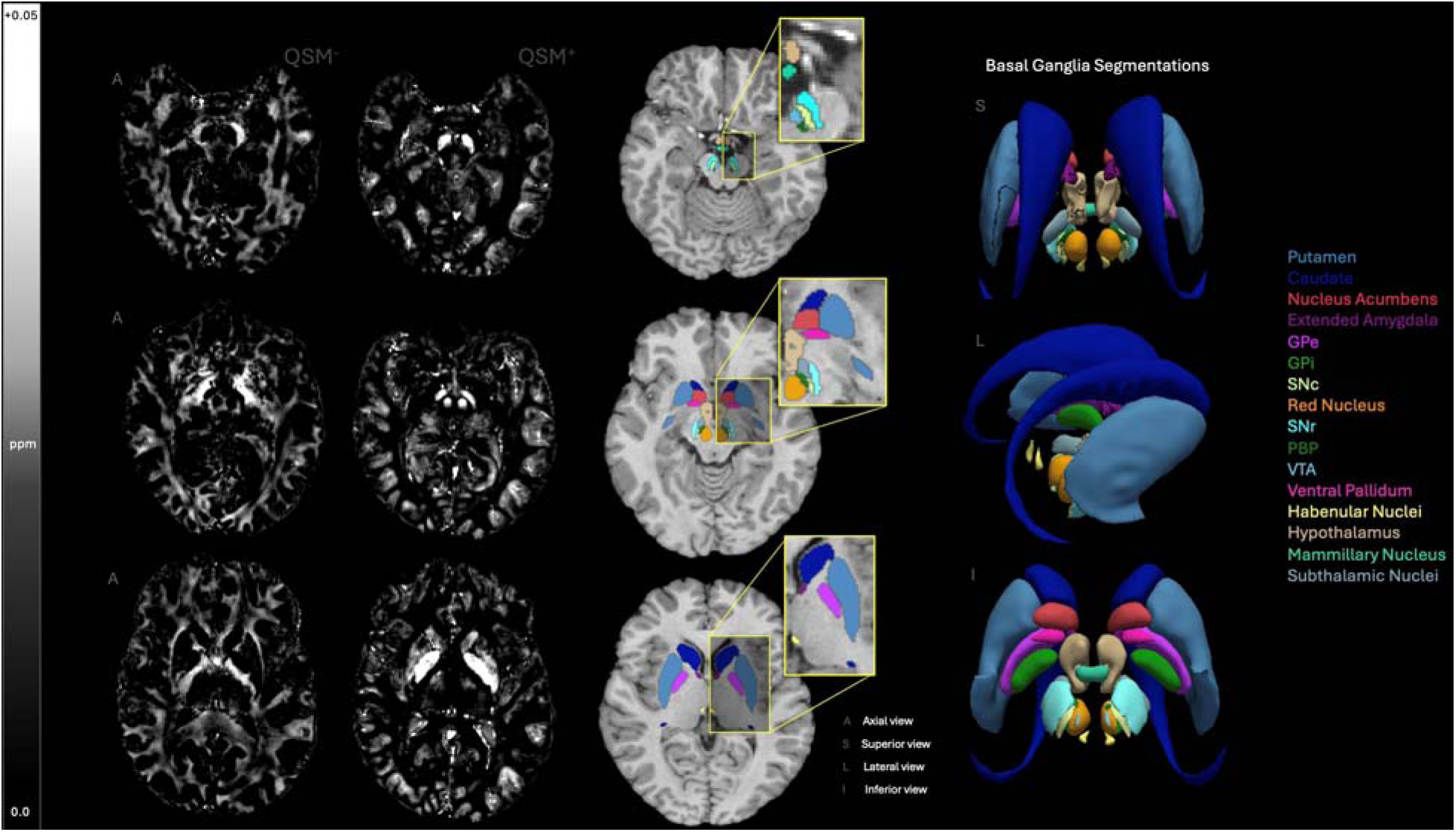
Segmentations of the basal ganglia. Sixteen bilateral segmentations of the basal ganglia and associated structures are displayed across axial (A) slices of the *T*_1_w CIT168-MNI152 brain template, as well as 3D renderings from superior (S), inferior (I), and lateral (L) perspectives for both the CIT168-MNI152 brain template and corresponding basal ganglia mask. The key for each deep grey nuclei is provided in the respective colour of that region. Axial slices for signed maps (*QSM*^−^ and *QSM*^+^) are shown at comparable depths for visual reference. Susceptibility values are expressed in parts per million (ppm) ranging from 0.0 to +0.05, and negative maps are multiplied by -1 for better visualisation. GPe = globus pallidus externus; GPi = globus pallidus internus; SNc = substantia nigra pars compacta; SNr = substantia nigra pars reticulata; PBP = parabrachial pigmented nucleus; VTA = ventral tegmental area.

#### Hippocampal segmentation

Prior to hippocampal segmentation, bias field correction was applied to each participants’ T_1_w images using the N4 algorithm^86^ from the Advanced Normalisation Tools (ANTs) library.^87^ The bias field-corrected T_1_w images were then processed with FreeSurfer’s recon-all pipeline.^88^ Unilateral segmentation of the hippocampal subregions was conducted using an automated, FreeSurfer-based pipeline^89^ built from a leverages a probabilistic atlas derived from ultrahighfield, ex vivo MRI data with approximately 0.1 mm isotropic voxel resolution. Of the three segmentations available, the mid-detail scheme “CA” scheme was selected for use in statistical analysis, balancing the level of segmentation detail with the need to mitigate the multiple comparisons problem. The hippocampal subregion masks included in this segmentation: the subicular complex (parasubiculum, presubiculum, subiculum), hippocampal subfields (including cornu ammonis [CA] regions CA1, CA3 [which includes CA2], and CA4), the hippocampal-amygdala transition area (HATA), fimbria, hippocampal tail, and hippocampal fissure. For full segmentation details, refer to Table 2, and for visual representation see Fig. 2.

**Table 2:** Hippocampal subregions and their closest anatomical structures in “CA” segmentation.

|  |  |
| --- | --- |
| Parasubiculum | Parasubiculum |
| Presubiculum head | Presubiculum |
| Presubiculum body |  |
| Subiculum head | Subiculum |
| Subiculum body |  |
| CA1 head | CA1 |
| CA1 body |  |
| CA3 head | CA3 |
| CA3 body |  |
| CA4 head | CA4 |
| CA4 body |  |
| GC-ML-DG head |  |
| GC-ML-DG body |  |
| Molecular layer HP-head | Closest structure* |
| Molecular layer HP-body |  |
| HATA | HATA |
| Fimbria | Fimbria |
| Hippocampal tail | Hippocampal tail |
| hippocampal fissure | hippocampal fissure |

\* = voxels in the molecular layer are assigned the label of the closest voxel that is neither molecular layer nor background. *Note.* This table details the unilateral segmentations of hippocampal subregions within the volume labelled as "CA", along with associated hippocampal structures. Subfield CA2 is incorporated into CA3. Table is also available at the FreeSurfer Wiki.

**Fig 2:**
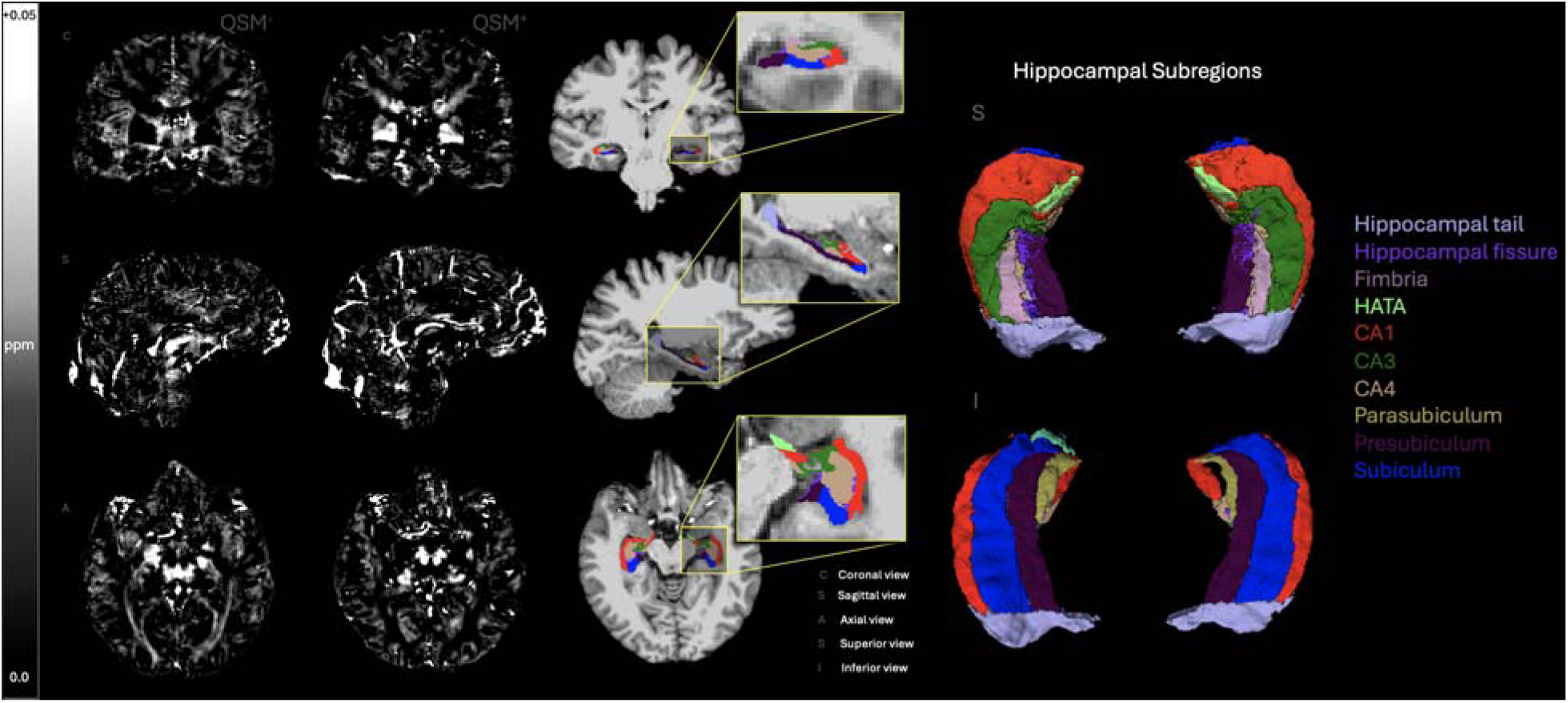
Segmentations of the hippocampal formation. Visual representation of the unilateral hippocampal segmentations provided by FreeSurfer under the “CA” scheme, displayed across coronal (C), axial (A), and sagittal (S) views as well as a 3D rendering from the inferior (I) and superior (S) views. The key for each hippocampal subregion is provided in the respective colour of that region. Signed maps (*QSM*^−^ and *QSM*^+^) from which susceptibility values are sampled for each hippocampal region are shown for each orientation (C, A, S). Susceptibility values are expressed in ppm ranging from 0.0 to +0.05, and negative maps are multiplied by -1 for better visualisation. HATA = hippocampal-amygdala transition area; CA 1-4 = cornu ammonis regions 1-4 (Note: CA2 is included in CA3).

To standardise each of the hippocampal subregion masks for data analysis, each subjects’ skull-stripped *T*_1_w brain image and left and right hemisphere hippocampal masks were converted from FreeSurfer’s .mgz format to NIfTI format (.nii.gz) using MRtrix3 *mrconvert*.^90^ After format conversion, each subjects’ *T*_1_w brain image was registered to the CIT168 *T*_1_w template in MNI152 space^79^ using *FLIRT*^80–82^ with 12 DoF. The resulting transformation matrix was then applied to both the left and right hemisphere hippocampal masks using nearest-neighbour interpolation to maintain the integrity of discrete hippocampal label boundaries during transformation. These steps ensured spatial alignment between each of the hippocampal masks, the signed susceptibility maps, and the CIT168 segmentations of the basal ganglia and associated nuclei provided in MNI152 space.^79^

### Statistical analyses

Statistical analyses were conducted at the bilateral level using MATLAB (2024a). Average positive and negative susceptibility values from both *QSM*^+^ and *QSM*^−^ maps were extracted from the 16 bilateral segmentations of the basal ganglia. For the 10 hippocampal regions, left and right hemisphere susceptibility values were extracted and averaged to yield a bilateral measure. Any ROIs in which zero values were present for any participant were omitted from subsequent analysis (the PBP for *QSM*^−^ analyses, and the HN for *QSM*^+^ analyses). There were no zero values for any participants for any hippocampal ROIs. To evaluate the effects of sr-mTBI compared to healthy controls, two-tailed independent sample t-tests were performed for each subcortical ROI, assessing group differences in susceptibility values. To control for multiple comparisons, a Benjamini-Hochberg false discovery rate (FDR) correction^91^ was applied separately to the 15 p-values corresponding to the remaining basal ganglia and associated nuclei (all nuclei except the PBP for *QSM*^−^ and HN for *QSM*^+^) and the 10 p-values for the hippocampal regions, for each signed susceptibility map. Since participants were precisely age-matched, age was not treated as a covariate or confounding factor in the between-group comparisons. However, to examine the associations between QSM values and age across the entire cohort, partial Pearson correlation coefficients were computed. These analyses assessed the potential linear relationship between age and both positive and negative susceptibility values across all subcortical ROIs, controlling for group status. Additional relationships were explored between susceptibility values and sr-mTBI-related variables using Pearson correlation coefficients, including BIST^68^ scores and injury latency (days since injury; DSI) variables, for clinical participants only. Due to missing BIST^68^ data, mTBI-17 was excluded from analyses related to injury severity. Correlations were also corrected for 15 and 10 ROI-wise comparisons, respectively, using FDR procedures.^91^

## Results

### Between-group differences

Results showed no significant differences in positive or negative susceptibility between sr-mTBI participants and HC for any of the basal ganglia and associated nuclei (*q >* 0.05; see Fig. 3).

**Fig 3:**
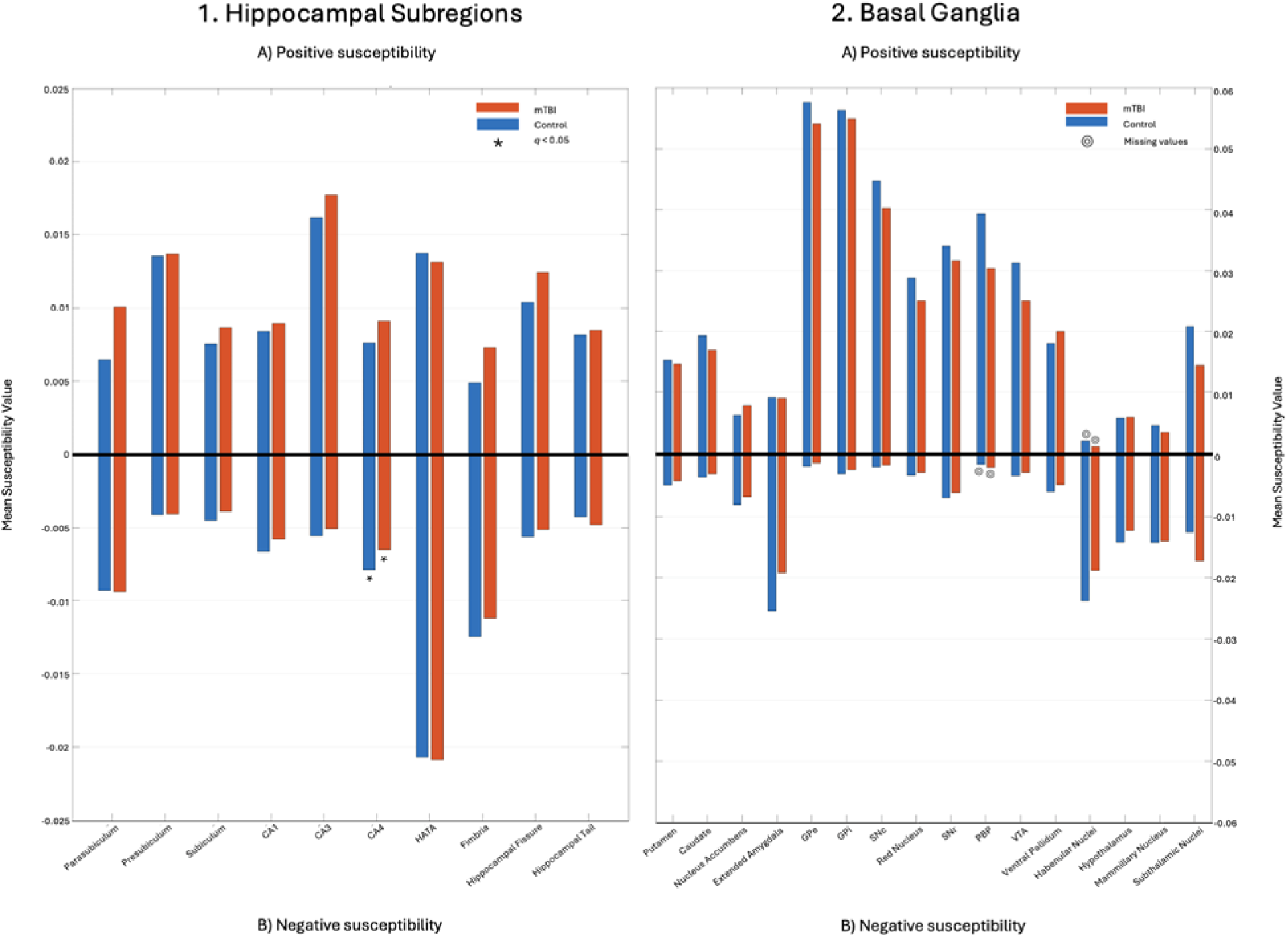
Mean susceptibility values in the hippocampal and basal ganglia ROIs. Bar plots showing the mean susceptibility values for mTBI (red bars) versus HC (blue bars) for: 1. Hippocampal subregions, and; 2. Basal ganglia and associated nuclei, for both A) positive and B) negative susceptibility maps. Susceptibility is measured in ppm. * = significant differences between groups; ⊙ = regions omitted from further analysis due to zero ROI-wise values for any participant.

There were no significant differences in positive susceptibility between groups for any of the 10 hippocampal subregions (see Fig. 3). Negative susceptibility values were significantly less negative for sr-mTBI participants (*M* = -0.007, *SD* = 0.002) than HC (*M* = -0.008, *SD* = 0.001) in the CA4 subfield only (*t*(48) = -2.99, *q* = 0.04). No significant differences between groups for negative susceptibility values were observed for any other hippocampal ROI.

### Age and susceptibility

No statistically significant correlations were observed between age and positive susceptibility values for any hippocampal ROI, however, a significant positive correlation between age and absolute negative susceptibility in the fimbria (*r*(48) = 0.42, *q* = 0.03) was observed, suggesting age-related increases in negative susceptibility (i.e., susceptibility becomes more negative with age (see Fig. 4)

**Fig 4:**
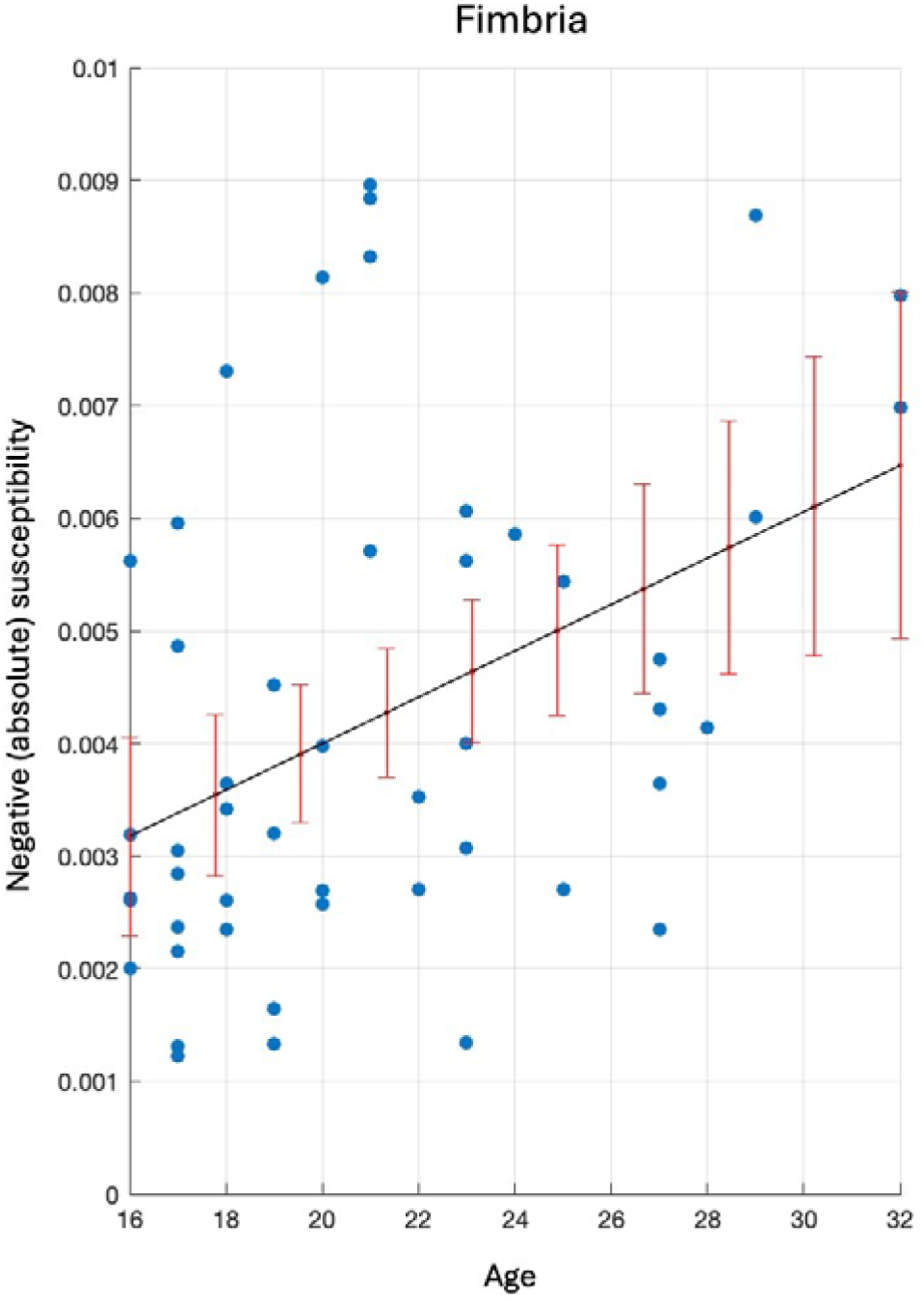
Significant correlations: Age and negative susceptibility in the fimbria. Scatter plot illustrating the statistically significant correlation between negative (absolute) susceptibility values in the hippocampal fimbria and age. A line of best fit (black) highlights the trend. 95% confidence interval bars (red) indicate the predicted range for true values, providing a measure of uncertainty in the model.

Results indicated significant age-related increases in positive susceptibility values in several basal ganglia or associated regions, including the putamen (*r*(48) = 0.64, *q <* 0.001), caudate (*r*(48) = 0.50, *q* = 0.001), red nucleus (*r*(48) = 0.66, *q <* 0.001), PBP (*r*(48) = 0.40, *q <* 0.02), and ventral pallidum (*r*(48) = 0.35, *q* = 0.04) (see Fig. 5).

**Fig 5:**
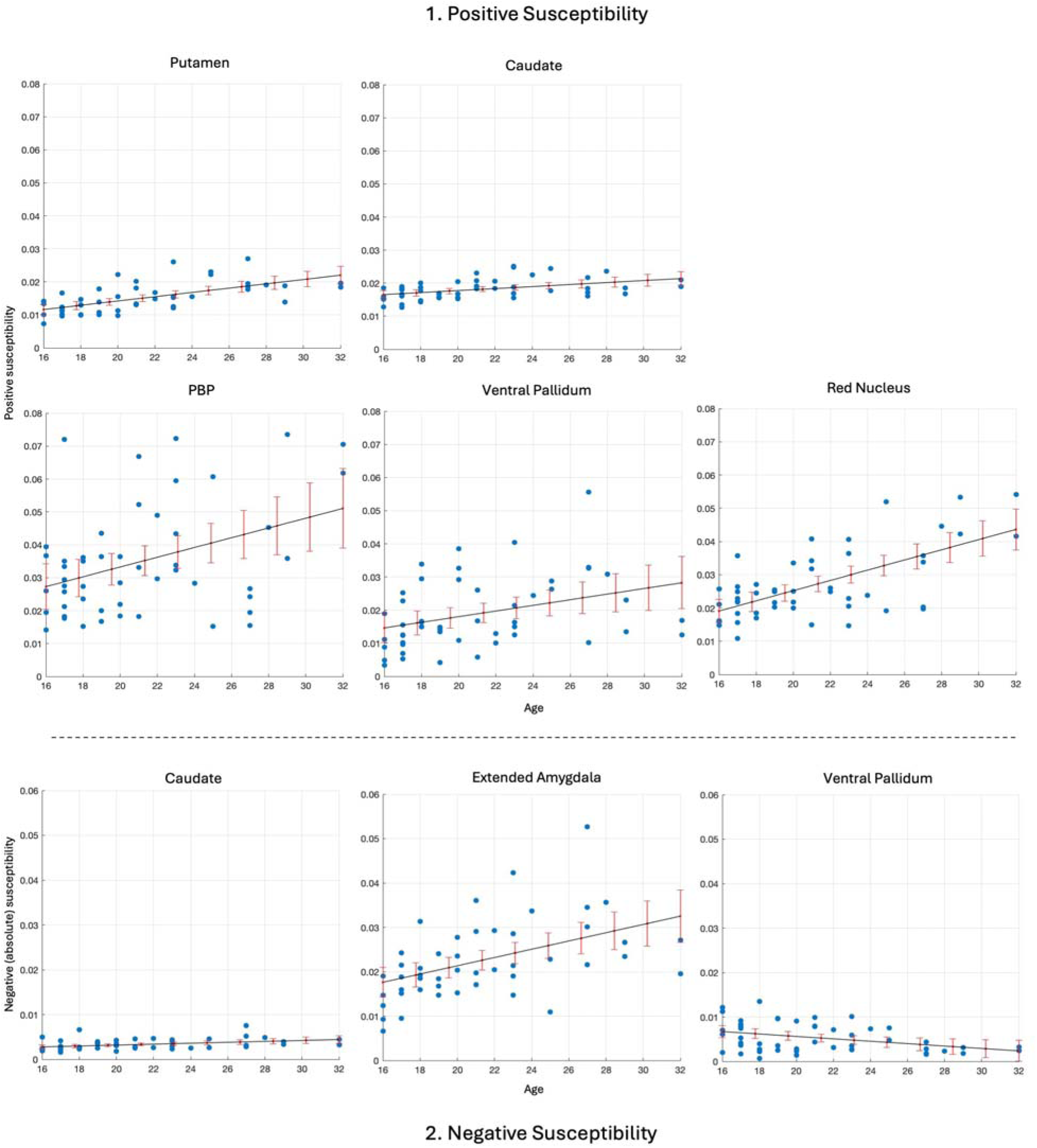
Significant correlations: Age and susceptibility values in the basal ganglia and associated nuclei. Scatter plots illustrating the statistically significant correlations between both 1. Positive and 2. Negative (absolute) susceptibility values and age in basal ganglia ROIs. A line of best fit (black) highlights the trend. 95% confidence interval bars (red) indicate the predicted range for true values, providing a measure of uncertainty in the model.

Significant positive correlations were also evident between age and absolute negative susceptibility in the caudate (*r*(48) = 0.39, *q* = 0.04) and extended amygdala (*r*(48) = 0.51, *q* = 0.002) suggesting that susceptibility in these regions tends to increase (become more negative) with age. Significant negative correlations were also observed between age and absolute negative susceptibility values in the ventral pallidum (*r*(48) = -0.37, *q* = 0.04), suggesting that susceptibility tends to decrease (become less negative) with age (see Fig. 5).

### Injury severity, injury latency, and susceptibility

No statistically significant correlations were observed between both positive and absolute negative susceptibility values and either DSI or BIST scores, for any basal ganglia and associated nuclei or hippocampal subregion.

## Discussion

Despite the vulnerability of the striatum and other deep nuclei to mechanical strain, cytoskeletal damage, and secondary metabolic disruptions after mTBI, all of which are risk factors for iron accumulation, previous QSM research has not yet achieved the anatomical specificity necessary to extensively characterise mTBI-related alterations to tissue content in the subcortical grey matter. To address this limitation, we performed a highly detailed segmentation of the basal ganglia and associated nuclei alongside the first QSM analysis of the hippocampal subregions in mTBI. QSM images were thresholded for inter-voxel sign prior to analysis. Between-group comparisons showed no significant differences between mTBI participants and controls for either susceptibility sign in the basal ganglia and associated nuclei, or for positive sign in the hippocampal subregions. However, the CA4 subfield exhibited significantly decreased (less negative) negative susceptibility values for the mTBI group relative to healthy controls. Correlational analyses also revealed no significant associations between either susceptibility sign and injury latency or severity in the mTBI group within any ROI. Congruent with expected age-related trends, positive susceptibility values in the putamen, caudate, PBP, ventral pallidum, and red nucleus increased with age. Age-related increases in negative susceptibility were also apparent in the fimbria, the caudate, and the extended amygdala, but decreased with age in the ventral pallidum.

### Negative susceptibility is decreased in CA4 subfield after mTBI

The observed decreased negative susceptibility in CA4 subfield reported here supports an axonal-injury model characteristic of both TBI^92^ and mTBI.^93^ Diffuse axonal injury is associated with disruptions of the myelin sheath^94,95^ or demyelination as demonstrated in both animal models^96^ and human QSM studies investigating sr-mTBI.^97^ Although density of myelinated fibers is relatively low in the hilus (CA4),^98^ disruptions to myelination in this region could plausibly account for localised decreases in negative susceptibility, suggesting this area could be vulnerable to axonal damage during mTBI.

Other plausible explanations for decreased negative susceptibility after mTBI could be related to phagocytosis of stressed cells and their components, including myelin. Microglia and macrophages are known to target and phagocytose distressed but still viable cells, even in the absence of cell death.^99^ This could result in a reduction in negative susceptibility if cells within a ROI are particularly sensitive to even mild injury and resultant distress signalling. Indeed, research suggests that the CA4 subfield is selectively vulnerable to sustained cell loss after head injury.^100,101^ Of particular relevance to this hypothesis, the hilus contains glutamatergic mossy cells and GABA-ergic somatostatinexpressing hilar interneurons which are highly vulnerable to damage and excitotoxicity in temporal lobe epilepsy, TBI, and mTBI.^102–104^ Indeed, murine models demonstrate irreversible loss of somatostatin-expressing hilar interneurons within just four hours of injury.^100^

In addition, mossy cells, unique to the hilus, are especially predisposed to metabolic disruptions and excitotoxicity, as well as premature neurodegenerative processes.^104^ Murine models also demonstrate the link between TBI and damage to hilar mossy cells, the “irritable mossy cell” hypothesis, which may account for the memory-related impairments observed after TBI.^103,104^ Mossy cells also express proteins that may be implicated in depression, and reductions in mossy cells may increase levels of anxiety^104^; both of which are mood-related symptoms of mTBI.^105^ Whilst speculation is interesting, in the absence of histological analysis, the precise pathological mechanisms cannot be accurately characterised. However, at minimum the results presented here support prior research noting the vulnerability of the hippocampus, and in particular the CA4 subfield, to mTBI-related pathology. Given the susceptibility of the CA4 subfield to both tau pathology in CTE^59,106^ and associated local dendritic swelling which are considered supporting features for diagnosis,^58,107^ alterations to neurites or vulnerable cell populations could speculatively represent an acute component of the injury cascade that culminates in tauopathy as a downstream pathological event later in life for some people.

### The effects of mTBI on positive susceptibility

This, and other,^41–47^ QSM investigations have found no significant evidence of iron accumulation in the basal ganglia or hippocampus after mTBI. However, iron accumulation as a pathologic feature of mTBI in many of these regions is well documented, contradicting the lack of iron-related findings reported here. For example, an iron-sensitive magnetic field correlation (MFC) study^37^ reported iron accumulation in the globus pallidus and thalamus following mTBI. Investigations using susceptibility weighted imaging (SWI) have also identified iron deposition in the caudate, thalamus, right substantia nigra, red nucleus, lenticular nucleus, splenium of the corpus callosum, and hippocampus in chronic mTBI, concomitantly linking accumulations in the substantia nigra with cognitive dysfunction.^54^ Studies using T_2_* imaging have identified iron deposition in cortical, subcortical (left putamen, bilateral hippocampal), and brainstem regions,^108^ along with reports of accumulation in the lateral geniculate nucleus of the thalamus,^109^ with both studies citing associations between elevated iron and post-mTBI headache symptomatology.

Additionally, animal models of controlled cortical impact TBI have demonstrated iron deposition in the thalamus ipsilateral to the impact site upon histological examination, with focal depositions co-localising *T*_2_ hypointensity on MRI.^110^ Murine models also report the vulnerability of the hippocampus to oxidative stress and synaptic protein modification, underscoring not only the regional susceptibility to traumatic injury mechanisms, but also highlighting the role of iron in the generation of cytotoxic free radicals.^111^ Post-mortem studies have provided evidence of iron accumulation in hippocampal neurofibrillary tangles (NFTs) in CTE,^61^ as well as abnormal haemosiderin-laden macrophages near small vessels in the frontal and temporal lobes less than one year post-injury, indicating ferritin bound non-haem iron deposition^112^ which is corroborated by recent research noting the presence of L-ferritin-positive astrocytes proximal to CTE lesions.^113^

Despite these studies highlighting the vulnerability of the basal ganglia and hippocampus to iron-mediated pathology at multiple stages post-injury, an absence of significant group differences in positive susceptibility observed in this study may suggest a general protective effect conferred by the deep subcortical location of these regions. In contrast, our prior work has demonstrated that the cortex, particularly the parahippocampal gyrus, is susceptible to iron-mediated pathology following mTBI.^48^ In more targeted, individualised analyses, cortical iron pathology has been observed in one-third of mTBI participants (rising to 83% when accounting for cortical depth and curvature) across various cortical ROIs, which was related to more severe injury-related symptoms.^114^

It is also reasonable to suggest that the absence of between-group differences in iron markers may reflect the influence of normal age-related increases in iron (for a review, see Madden and Merenstein^17^), which could obscure subtle mTBI-specific effects. Age-related iron deposition is particularly pronounced in the basal ganglia and most evident within the age range of participants in this study, as discussed in more detail further below. This shared characteristic among younger individuals may explain the lack of significant group differences in iron-related markers. In contrast, the general lack of age effects in hippocampal regions and the resultant sensitivity to disruptions in negative susceptibility in the CA4 subfield supports the hypothesis that changes to biomagnetism which are common to all participants may obscure injury-related susceptibility effects. Consequently, the unique alteration to negative susceptibility values in the CA4 subfield thus likely reflect specific pathological consequences of mTBI.

In keeping with these observations, thresholding bulk magnetic susceptibility likely enabled the isolation and identification of weaker negative susceptibility effects, which could otherwise have been diluted or overshadowed by stronger positive susceptibility signals during analysis.^26^ This could also account for the lack of significant differences reported in previous studies where neither thresholding or magnetic source separation techniques suitable for multi-echo data were employed to generate separate sign-wise maps,^41–47^ providing support for intra- or inter-voxel susceptibility separation techniques, depending on the data acquisition parameters.

### The effects of mTBI on negative susceptibility

We found no evidence of increased negative susceptibility in the subcortical grey matter following mTBI. However, alterations to varying biomagnetic substrates following TBI have previously been characterised in murine models using QSM and R * imaging alongside histological examinations to assess changes not only to iron, but also myelin and calcium.^115^ These models have demonstrated concurrent iron and calcium accumulation, as well as demyelination after impact. These findings are reinforced by additional *in vivo* and *ex vivo* murine models of TBI also reporting significant calcifications, which are proposed as an indirect measure of inflammation^116^ and microglia activation^117^ following injury. Of particular interest, studies have noted that calcifications often co-localise with paramagnetic iron in TBI.^115^ This finding underscores the complex interplay of tissue components affected by mTBI and highlights the contribution of both paramagnetic and diamagnetic substrates to injury pathology. This further validates the need for either source separation, if data are acquired with multi-echo sequences,^118–123^ or single-echo-compatible voxel-wise thresholding,^83,84^ in QSM research to differentiate these overlapping effects and better elucidate underlying biological mechanisms.

A*β* and tau, two proteins with potentially diamagnetic properties,^31–33^ have been associated with both acute injury pathology and later neurodegenerative processes, and are known to co-localise with iron in Alzheimer’s disease (AD)^17^ and CTE.^61^ Focal accumulations may be present in the entorhinal cortex, hippocampus, and striatum (caudate and putamen) as evidenced by PET studies of TBI.^124^ The aggregation of both proteins would likely produce differences in negative susceptibility on QSM. However, systematic reviews have noted inconsistencies between studies regarding the presence or absence of Aβ as a pathologic feature of TBI,^124^ and while sulcal p-tau lesions are diagnostic of CTE, it remains challenging to identify deposition *in vivo* and thus assess temporal dynamics.^125^ These factors may have contributed to the absence of quantifiable increases in negative susceptibility in the present study. Alternatively, and perhaps more plausibly, the results presented here indicate that A*β* and tau are not present in this young cohort^126,127^ imaged at the acute stage of a mild head injury.

### Temporal biodynamics of positive susceptibility in ageing

Iron increases throughout the lifespan as a function of normal ageing.^17,63^ Particularly steep increases in brain iron content are present in the red nucleus and substantia nigra within the first 20 years of life, the globus pallidus at approximately 30 years, with maximal values reached in the putamen and caudate at approximately five to six decades after birth.^35^ This histological study is supported by extensive QSM investigations,^17^ for example, QSM and R * mapping have demonstrated susceptibility increases in the putamen and globus pallidus throughout adulthood, with values in the caudate peaking in the third decade.^66^ Other QSM studies have evidenced increased susceptibility values in the caudate, putamen, globus pallidus, red nucleus, and substantia nigra in healthy older populations relative to younger samples.^64^ Both imaging and histological studies support the biological plausibility of correlational results presented here, particularly within the caudate, putamen, and red nucleus.^17^ This study also extends the existing literature through the observation of additional age-related positive susceptibility increases in the PBP and ventral pallidum. Our correlational findings, and evidence from the literature more broadly, suggest that a younger cohort is particularly prone to iron deposition as a shared characteristic, irrespective of mTBI status. For hippocampal regions, on the other hand, age-related iron accumulation apparent in previous research^17,63,65,67^ was not reflected in the results of this study, potentially due to a likelihood of more subtle effects in these regions, coupled with the limited age range of this sample.

### Temporal biodynamics of negative susceptibility in ageing

The temporal dynamics of diamagnetic substrates are relatively under-investigated in QSM, making inferences far less substantive than those related to paramagnetism and iron. Although myelin is the main source of negative susceptibility on QSM^27^ it is unlikely that increases in negative susceptibility reflect age-related increases in myelin content. Whilst developmental myelination of axons is apparent up until approximately 30 years of age, this protraction of normal processes is generally restricted to prefrontal cortical regions.^128^ In other brain regions, steep increases in myelin content are most apparent in the first months of life, after which time any disparities between age groups are unlikely to produce noticeable differences in tissue contrast on MRI.^129^ Age-related kinetics of Aβ and tau aggregation point toward similar conclusions. While deposits of Aβ have been detected in the brains of individuals as young as 20 years old,^130^ it is typically a feature associated with older age, and has been noted in both the presence and absence of cognitive dysfunction^126^; a similar pattern to that observed in tau accumulation.^127^ As such, trends for increasing negative susceptibility with age are unlikely to be related to protein accumulation. Here, reasonable speculation suggests that the positive relationship between negative susceptibility and age in the caudate, extended amygdala, and fimbria may be related instead to calcifications in these regions. This is supported by a body of work highlighting age-related calcifications in the basal ganglia^131,132^ and hippocampus^133^ in healthy individuals. However, the paucity of granular segmentations in the wider literature makes drawing parallels with the present research more difficult.

Conversely, the decrease in negative susceptibility as a function of age in the ventral pallidum may be related to myelin changes. The ventral pallidum contains both neuronal cell bodies and myelinated axons for rapid signal transmission.^134–136^ Although literature related to age-induced demyelination specific to this region does not abound, investigations of myelin biodynamics more broadly suggests a 10% decrease per decade between the ages of 20 and 80,^137^ which may account for the moderate effects evident here.

### Injury latency and severity do not affect tissue magnetic susceptibility

While research suggests a negative correlation between iron deposition and cognitive ability following mTBI,^54^ self-reported injury severity is often misaligned with objective markers of pathology, including on neuroimaging.^138,139^ Alongside the under-reporting of symptoms that often occurs following a sr-mTBI,^140,141^ these factors could confound results and contribute to the lack of correlation between positive or negative susceptibilities and both BIST scores and injury latency variables observed here. However, results from a previous investigation have suggested a link between injury severity and *cortical* iron dyshomeostasis.^114^ The cortex is particularly vulnerable to a diverse range of injury mechanisms and downstream pathology in mTBI, including acute mechanical deformation and sulcal injury, which may be implicated in cortical atrophy and tauopathy in CTE.^60,142–145^ It is therefore not unreasonable to suggest that cortical, rather than subcortical, brain tissue content changes may be more predictive of adverse symptomatology after mTBI. Finally, the lack of correlation between positive or negative susceptibilities and injury latency suggests that mTBI-related neuropathology may be time-dependent and more evident at subacute and chronic stages. Additional longitudinal research may be needed to better represent associations between injury latency and alterations to brain tissue content.

### Limitations

The absence of elevations in positive or negative net magnetic susceptibility following mTBI suggest that iron deposition, calcifications, or focal aggregation of proteins such as tau and Aβ may not be present at the acute stage of injury. However, aforementioned research provides strong evidence that head injury more broadly, and even mild instances, can lead to diverse changes across biomagnetic substrates. Several factors may have inhibited the detection of changes in this research. Firstly, focal accumulations of iron, calcium, or proteins may better characterise chronic or later-life effects rather than acute pathophysiology, as evidenced by neuroimaging studies where differences in iron deposition were apparent at an average of 559 days post-injury.^37^ Secondly, tau elevations are most commonly reported in cases of repeated exposure to head injuries,^125^ and in the absence of prior injury data, the study design precluded stratification of mTBI participants according to exposure. Without this information, detection of negative susceptibility changes and the potential relationship with tau pathology may be inhibited. Future studies should consider longitudinal research designs tracking athletes’ exposure to head injury to better elucidate the cumulative and temporal effect of mTBI on biomagnetic substrates.

Notwithstanding the detail of the segmentations used in this study relative to previous more macroscopic investigations, the spatial resolution of the susceptibility maps may still limit detection of microstructural alterations. Because the susceptibility value at each voxel in a QSM image is obtained by convolving the susceptibilities of neighbouring voxels with a dipole kernel, each voxels susceptibility value is influenced by the surrounding susceptibility distribution,^146^ and thus resolutions of 1 mm and above may hinder the detection of the subtle pathophysiological changes associated with mTBI. However, it should be noted that a 1 mm isotropic voxel resolution still meets the minimum criteria for best-practice QSM.^77^ Additionally, the use of single-echo QSM restricted voxel thresholding to an inter-voxel approach. As a result, the presence of multiple biological substrates with opposing magnetic properties *within* voxels may have confounded results by representing an aggregate of all susceptibility sources. These bipolar contributions within voxels cannot be disambiguated, and thus analysed, without multi-echo acquisitions.^118–123^ This was also a likely cause of missing values in certain ROIs comprised primarily of either positive or negative susceptibility sources, such as the habenular nuclei or PBP (see Fig. 3). Thresholding these extremely polarised regions may result in zero values for the nondominant sign for some participants, which may be further exacerbated by the size of ROIs, like the relatively small habenular nuclei. The use of multi-echo acquisitions would likely also resolve this limitation. In addition, group-level analyses may obscure individual neuropathology and inter-individual heterogeneity^114,147,148^ and less macroscopic statistical approaches are currently planned for future research.

While the hippocampal^89^ and basal ganglia^79^ atlases offer robust segmentations, several limitations are associated with these approaches. First, the hippocampal segmentations provided via FreeSurfer^88^ were developed from a small number of donors (10 for healthy controls, up to 15 when including mild cognitive impairment and AD subjects), with advanced age at death (60-91 years), which may result in hippocampal atrophy influencing the delineations. Furthermore, even with ultrahigh resolution MRI, some regional boundaries are not clearly visible in the training data. Similarly, for the basal ganglia atlas, some fine anatomical details may be lost, potentially leading to inaccuracies in boundary delineation. Although manual delineation is considered the gold standard, it requires extensive anatomical expertise and is prohibitively time-consuming.^149^ Recent advances in deep-learning-based hippocampal segmentation have shown promise^150^; however, given that the current cohort is unlikely to exhibit gross hippocampal atrophy or other dramatic structural alterations, these advanced approaches may be more beneficial in samples where such abnormalities are expected.

Lastly, this research was conducted on a sample exclusively comprised of males, limiting external validity. This is particularly relevant to the translation of findings reported here to female sports players. Sex can influence time dependence of the neuroinflammatory response,^125^ and fluctuating sex hormone profiles,^151^ oral contraceptive use,^152^ and variability in neck muscle morphology^153^ can contribute to differences in injury severity and outcome. Further research is planned within this demographic, and will ensure generalisability to female athletes of this age range.

### Conclusions

This research builds upon previous QSM studies of mTBI by presenting the first assessment of mild injury effects specifically in the hippocampal subregions, and offers the most detailed segmentation of the basal ganglia and associated nuclei to date. Here, both positive (iron-related) and negative (myelin-, calcium-, and protein-related) net susceptibility maps were utilised to better understand the impact of mTBI on biological substrates with differing biomagnetic profiles. Results revealed mTBI-related decreased negative susceptibility in the CA4 subfield, indicating potential disruption to myelin or vulnerable cell populations in this region which may represent a component of the degenerative injury cascade. The absence of significant increases in either susceptibility sign as a marker of mTBI pathophysiology may reflect a general protective effect of subcortical location within the brain, or the confounding effect of age-related susceptibility changes common to all participants. Finally, results from correlational analyses support and extend prior literature regarding age-related iron deposition in subcortical grey matter and contributes to the sparse literature on the relationship between negative susceptibility values and age.

## Data Availability

All data produced in the present study are available upon reasonable request to the authors

## Author Contributions

**Christi A. Essex** (Conceptualisation, Methodology, Project Administration, Validation, Software, Formal Analysis, Investigation, Resources, Data Curation, Writing - Original Draft, Writing - Review & Editing, Visualisation); **Mayan J. Bedggood** (Writing - Review & Editing, Project administration, Investigation); **Jenna L. Merenstein** (Writing Review & Editing); **Helen Murray** (Writing - Review & Editing); **Catherine Morgan** (Methodology, Writing - Review & Editing); **Samantha J. Holdsworth** (Writing - Review & Editing); Richard L.M. Faull (Writing - Review & Editing); **Patria Hume** (Writing - Review & Editing); **Alice Theadom** (Conceptualisation, Methodology, Writing - Review & Editing, Funding acquisition, Supervision); **Mangor Pedersen** (Conceptualisation, Methodology, Writing Review & Editing, Funding acquisition, Supervision)

## Acknowledgements

We extend thanks to Amabelle Voice-Powell and Cassandra McGregor for their contribution to the data collection, and Tania Ka’ai for bringing her perspective to cultural considerations on this study. In addition, we thank Axis Sports Concussion Clinics, particularly Dr Stephen Kara, for their assistance with recruiting sr-mTBI participants and personnel at the Centre for Advanced Magnetic Resonance Imaging (CAMRI) for their assistance collecting MRI data. We also acknowledge Dr Tim Elliot for radiological reporting of all participants and Siemens Healthineers for the use of a work-in-progress (WIP) prototype sequence for the acquisition data used to perform QSM.

## Data availability

De-identified MRI data and code used for image processing and statistical analysis can be made available upon request to the corresponding author.

